# Provider Confidence in Counseling Preconception, Pregnant, and Postpartum Patients Regarding COVID-19 Vaccination

**DOI:** 10.1101/2021.12.17.21267998

**Authors:** Molly R. Siegel, Kaitlyn E. James, Elana Jaffe, Michelle M. L’Heureux, Anjali J. Kaimal, Ilona T. Goldfarb

**Affiliations:** Department of Obstetrics and Gynecology, Massachusetts General Hospital, Boston, MA; School of Medicine, University of North Carolina at Chapel Hill; Massachusetts General Hospital, Boston, MA

**Keywords:** Pregnancy, vaccine hesitancy, COVID-19, prenatal, postpartum, preconception, vaccine counseling

## Abstract

**Objective:** To assess knowledge and confidence in COVID-19 vaccine counseling among practitioners who provide care to pregnant persons and to describe factors associated with confidence in counseling.

**Study Design:** A web-based anonymous survey was distributed via email to a cross-sectional convenience sample of OB/GYN, Primary Care, and Internal Medicine faculty at three hospitals in a single healthcare network in Massachusetts. Individual demographics and institution-specific variables were included in the survey along with questions assessing both attitudes toward COVID-19 illness in pregnancy as well as confidence in counseling regarding the use of the vaccine for pregnant patients.

**Results:** Almost all providers (151, 98.1%) reported already receiving or being scheduled to receive a COVID-19 vaccine, and a majority of providers (111, 72.1%) reported that they believe the benefits of the mRNA COVID-19 vaccine in pregnancy outweigh the risks. Forty-one (26.6%) reported feeling very confident in counseling patients who primarily speak English about the evidence for mRNA vaccination in pregnancy, and 36 (23%) reported feeling very confident in counseling patients who are not primarily English-speaking. Forty-three providers (28.1%) expressed strong confidence in their comfort talking to individuals with vaccine hesitancy based on historic and continued racism and systemic injustices. The sources that survey respondents most commonly used to find information regarding COVID-19 vaccination in pregnancy were the CDC (112, 74.2%), hospital-specific resources (94, 62.3%) and ACOG (82, 54.3%).

**Conclusion:** While providers reported high personal uptake of COVID-19 vaccination and felt that the benefits of vaccination outweigh the risks in pregnancy, less than one-third felt very confident in counseling pregnant patients about available evidence for mRNA vaccine safety in pregnancy. Ensuring that providers feel comfortable bridging the gap between their belief that the vaccine is beneficial for pregnant patients and their comfort with holding conversations with patients regarding vaccination is paramount in order to ensure equitable access to vaccines for pregnant patients.

## Background

Pregnant people were considered a priority population for COVID-19 vaccination in a majority of states.^1^ Their prioritization in vaccine eligibility stems from growing evidence that pregnant individuals are at increased risk of complications from COVID-19 infection, including hospitalization, intensive care unit (ICU) admission, and need for invasive ventilation, and death.^2^ As with other groups at increased risk for complications due to COVID-19, they stand to benefit from vaccination. However, pregnant and lactating persons were not included in early trials of the vaccine, and as such, there are fewer data to inform use in these groups. As a result, patients face potentially challenging decisions surrounding vaccination in pregnancy and lactation. Together with their providers, pregnant individuals must consider the high level of protection afforded by the vaccine together with the limited evidence around vaccine safety in pregnancy, the established increased risk of morbidity and mortality associated with contracting the virus while pregnant, and their individual level of risk due to occupation, underlying medical conditions, or other factors.

Despite lack of data from randomized controlled trials, the Centers for Disease Control (CDC) and the Advisory Committee on Immunization Practices (ACIP), along with the American College of Obstetricians and Gynecologists (ACOG) and the Society for Maternal Fetal Medicine (SMFM), all support the provision of these vaccines to pregnant and breastfeeding individuals. While consultation is not required, these organizations encourage patients to seek counsel from their healthcare providers in deciding whether or not to receive the vaccine and recommend that healthcare providers engage in shared decision making with patients.^3,4^ As pregnant and lactating individuals consider whether they should get the COVID-19 vaccine, it is paramount that providers feel knowledgeable and comfortable counseling patients. However, healthcare provider knowledge and confidence surrounding COVID-19 vaccine counseling in pregnancy and lactation has not been assessed.

The purpose of this survey study is to assess knowledge and confidence in COVID-19 vaccine counseling among practitioners who provide care to pregnant patients and to describe factors associated with confidence with counseling. It is our goal that understanding these factors will help to identify resources needed to improve healthcare provider confidence and improve counseling around shared decision making for vaccination for pregnant individuals.

## Methods

### Participants

The survey participants were drawn from a cross-sectional convenience sample of OB/GYN, Primary Care, and Internal Medicine faculty at three hospitals (one urban academic medical center with 3800 deliveries per year, one suburban community hospital with 4500 deliveries per year, and one rural community hospital with 600 deliveries per year) in a single healthcare network in Massachusetts. Participants were asked via email to participate in an online anonymous survey in February 2021. The survey email was sent to MD, DO, CNM, and NP faculty by their division administrators with two follow-up reminder emails. Consent to participate in the survey was implied by voluntary participation in the anonymous survey.

### Survey

The web-based anonymous survey was designed to accommodate quantitative analysis of questions with nominal, ordinal, and interval level measurement and was reviewed by 3 experts in qualitative research methods and survey design. Individual demographics and institution-specific variables were included in the survey along with questions assessing both attitudes toward COVID-19 illness in pregnancy as well as confidence in counseling regarding the use of the vaccine for preconception, pregnant, and lactating patients.

The study and survey instrument were approved by the Partners Healthcare IRB (IRB # 2021P000088).

### Data Analysis

Data were analyzed using Stata/IC 16.1. For quantitative variables, descriptive statistics are presented to illustrate the distribution of the respondent demographics and survey responses. To determine predictors of confidence in counseling, multivariable logistic regression models were used. Separate models were generated to determine predictors of confidence and agreement (as defined as responding “very confident” or “strongly agree”) in counseling English-speaking and non-English speaking patients about mRNA COVID-19 vaccine safety, and in counseling patients with vaccine hesitancy stemming from historic and ongoing systemic racism and a belief that vaccines in general are harmful. Covariates for the multivariable models were chosen a priori; all models included provider specialty, frequency of being asked advice about COVID-19 vaccine, experience managing patients with COVID-19, and a personal belief that the COVID-19 vaccine benefits outweighed the risks. Confidence in counseling non-English speaking patients also included whether the respondent spoke more than one language; provider race was additionally included in the model for counseling patients with vaccine hesitancy based on historic and continued racism. P<0.05 was considered statistically significant.

## Results

### Provider demographics

There were 154 providers who responded to the survey. Demographic characteristics are shown in **Table 1**. Almost half of providers worked in obstetrics; approximately 70% of participants were medical doctors and a third were advanced practice practitioners. Most respondents identified as White/Caucasian and reported English as their primary language, while almost half of respondents reported speaking another language in addition to English.

**Table 1:** Survey Respondents

|  |  |
| --- | --- |
|  | N=154 |
| <u>Age (mean, SD)</u> | 46.2 (11.5) |
| <u>Race</u> |  |
| Black/African American | 6 (3.9%) |
| White | 131 (85.1%) |
| Asian | 11 (7.1%) |
| Other | 3 (1.9%) |
| Not reported | 3 (1.9%) |
| <u>Ethnicity</u> |  |
| Hispanic/Latinx | 5 (3.2%) |
| Not Hispanic/Latinx | 142 (92.2%) |
| Not reported | 7 (4.5%) |
| <u>Language</u> |  |
| English as primary language | 153 (99.4%) |
| At least one other language | 76 (49.4 %) |
| <u>Gender</u> |  |
| Female | 124 (80.5%) |
| Male | 24 (15.6%) |
| Non-binary | 1 (0.6%) |
| Prefer not to answer | 3 (1.9%) |
| Not reported | 2 (1.3%) |
| <u>Years in healthcare</u> |  |
| < 5 years | 7 (4.5%) |
| 5-10 years | 37 (24.0%) |
| 10-20 years | 38 (24.7%) |
| >20 years | 70 (45.5%) |
| Not reported | 2 (1.3%) |
| <u>Specialty</u> |  |
| OB-Only | 13 (8.4%) |
| OB/GYN | 62 (40.3%) |
| Primary Care | 46 (29.9%) |
| Internal Medicine | 21 (13.6%) |
| Other | 10 (6.5%) |
| Not reported | 2 (1.3%) |
| <u>Practitioner type</u> |  |
| MD/DO | 109 (70.8%) |
| CNM | 26 (16.9%) |
| NP | 19 (12.3%) |
| <u>Practice Setting</u> |  |
| Private practice, community-hospital | 10 (6.5%) |
| Group practice, community hospital | 48 (31.2%) |
| Private practice, academic hospital | 3 (1.9%) |
| Group practice, academic hospital | 86 (55.8%) |
| Other | 7 (4.5%) |

The majority of participants (140, 91.5%) reported that their institution was offering the COVID-19 vaccine to eligible pregnant individuals, and 137 (89.5%) reported that their institution was offering the vaccine to postpartum individuals. Most providers (126, 81.8%) reported that since December 2020, they had been asked to offer advice surrounding COVID-19 vaccination in pregnancy and postpartum to patients at least once per week, and slightly less than half (67, 43.5%) % participated in this counseling 2-10 times per week. Most providers (124, 80.5%) had cared for patients with COVID-19 in the outpatient setting, though fewer (73, 47.4%) had cared for pregnant outpatients with COVID-19. Similarly, while just over half of providers had managed patients with COVID-19 in an inpatient setting, fewer (58, 37.7%) reported managing pregnant patients with COVID-19 in the inpatient setting.

Survey respondents had high COVID-19 vaccine utilization for themselves. Almost all providers (151, 98.1%) reported already receiving or being scheduled to receive a COVID-19 vaccine. Respondents also reported that they frequently recommend other vaccines in pregnancy. Almost all providers (142, 92.2%) reported that they always recommend the flu vaccine for pregnant individuals during flu season, and the majority (119, 77.3%) reported that they recommend the Tdap vaccine for pregnant individuals during every pregnancy.

### Provider beliefs about risk-benefit ratio

Beliefs about the efficacy and safety of mRNA COVID-19 vaccines are shown in **Figure 1**. A majority of providers (111, 72.1%) reported the opinion that the benefits of the mRNA COVID-19 vaccine in pregnancy outweigh the risks. Several (21, 13.6%) were unsure about the risk-benefit ratio and agreed that additional data and more directive guidance from governing societies would be needed to guide a definitive opinion. A similar number (21, 13.6%) reported that the benefits outweighed the risks only in specific circumstances, including where patients had underlying co-morbidities, increased occupational exposure, lived in congregate housing, or were experiencing incarceration. Only 1 provider (0.6%) stated the belief that the benefits of the COVID-19 vaccine did not outweigh the risks of vaccination in pregnancy, citing unknown fetal safety. The most common rationales supporting the belief that the benefits of COVID-19 vaccines outweighed any risk were that pregnant individuals face increased risk of severe disease from COVID-19 infection, the likely low theoretical risk of mRNA COVID-19 vaccines in pregnancy, and because vaccine efficacy is likely to be similar between pregnant and non-pregnant individuals. The majority of providers (141, 92.2%) agreed that the benefits of COVID-19 vaccination during breastfeeding outweighed any risks, with 11 (7.2%) unsure and none in disagreement. Similarly, the majority of providers (133, 86.9%) agreed that the benefits of COVID-19 vaccines outweigh the risks for individuals planning pregnancy within three months.

**Figure 1:**
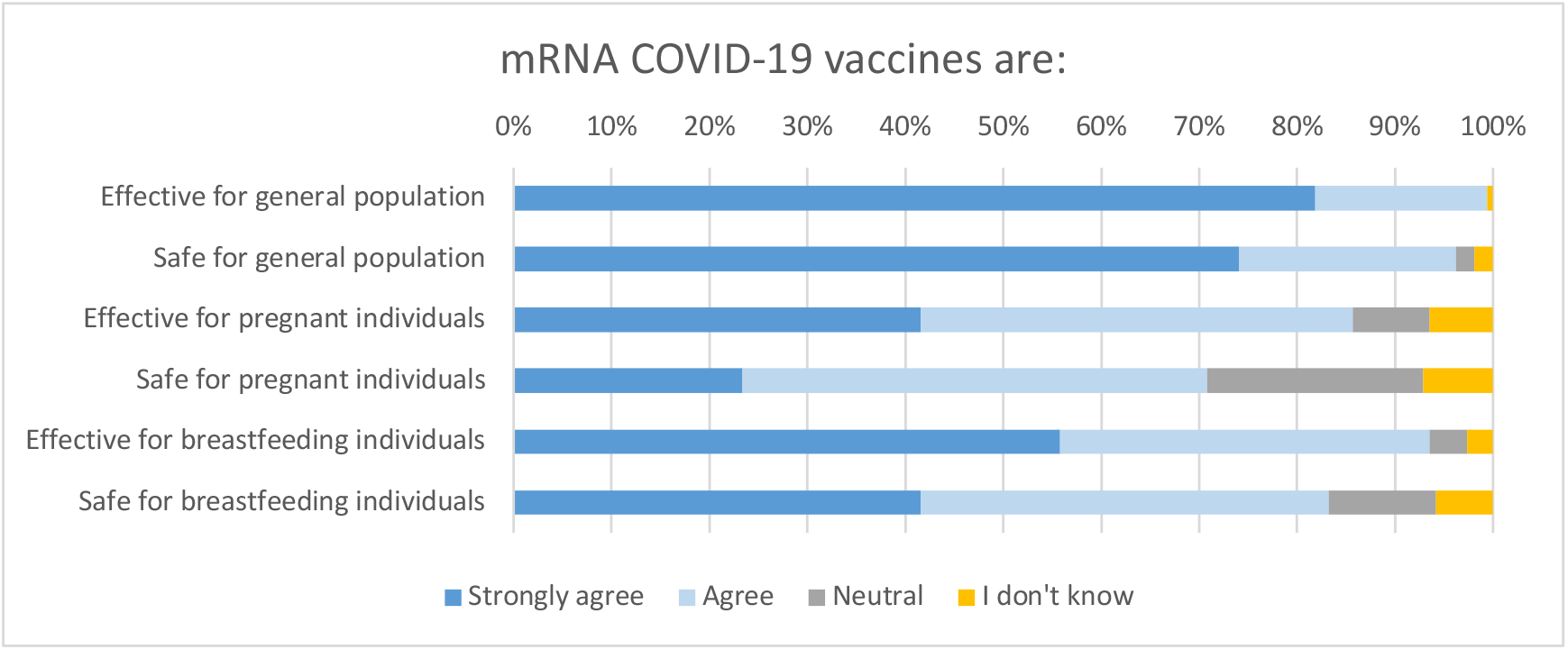
Attitudes towards mRNA COVID-19 Vaccine

Provider confidence in counseling patients who primarily speak English and those who do not primarily speak English about the vaccine is shown in **Figure 2**. When asked if that they could answer questions about the available evidence for mRNA COVID-19 vaccine safety in pregnant individuals, 41 (26.6%) reported feeling very confident in counseling patients who primarily speak English, and 36 (23%) reported feeling very confident in counseling patients who are not primarily English-speaking. There were no differences in confidence in counseling English-speaking patients about vaccine safety in pregnancy by provider type, but primary care and internal medicine providers were more likely to state that they feel very confident answering questions from non-English-speaking patients about the available evidence of mRNA COVID-19 vaccine safety in pregnant individuals (aOR 4.14, 95% CI 1.25-13.7).

**Figure 2:**
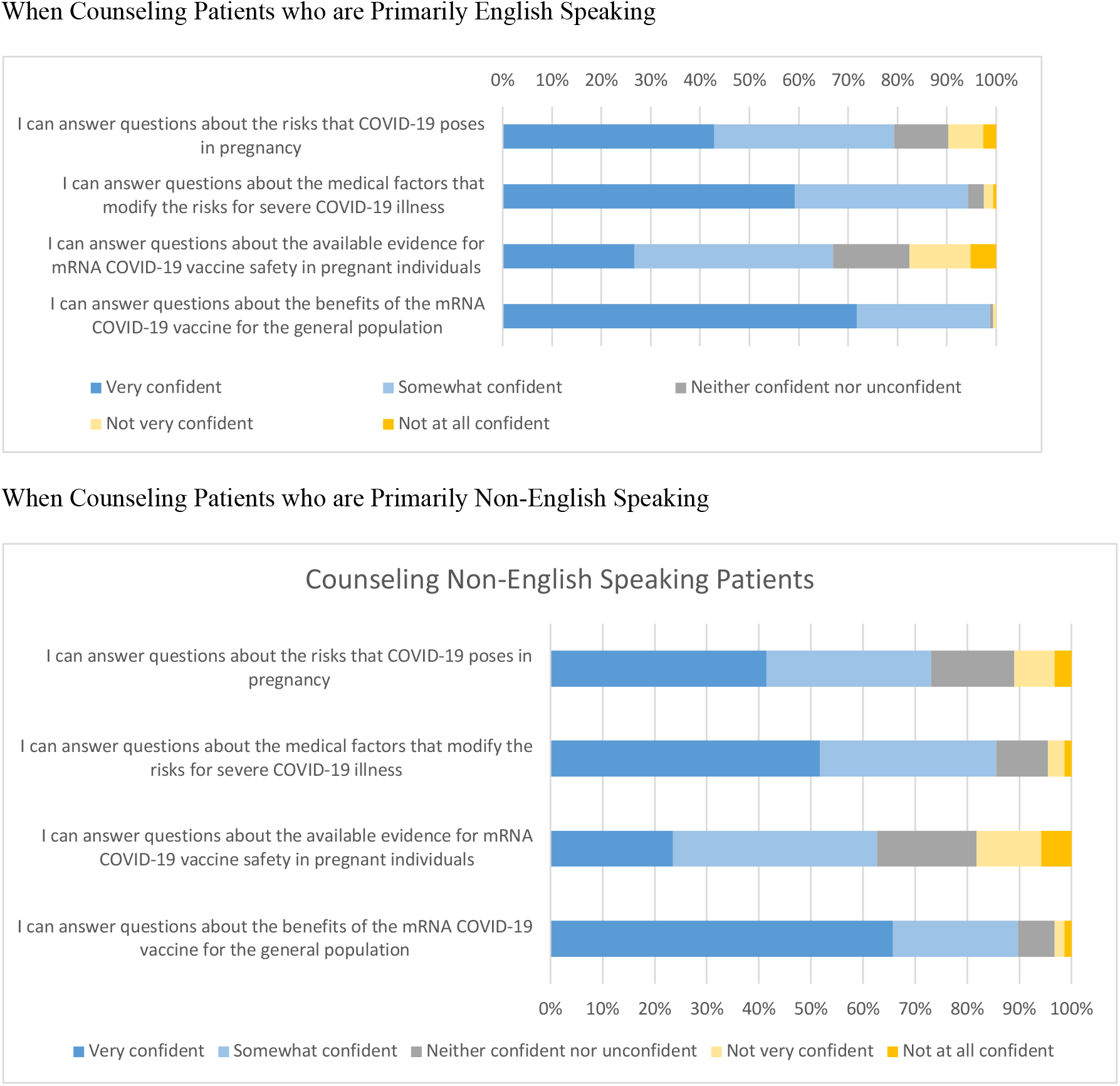
Counseling Confidence in Primarily English Speaking and Primarily Non-English-Speaking Patients

### Provider comfort with counseling patients with vaccine hesitancy

#### Provider comfort with counseling patients with vaccine hesitancy is shown in Figure 3

**Figure 3:**
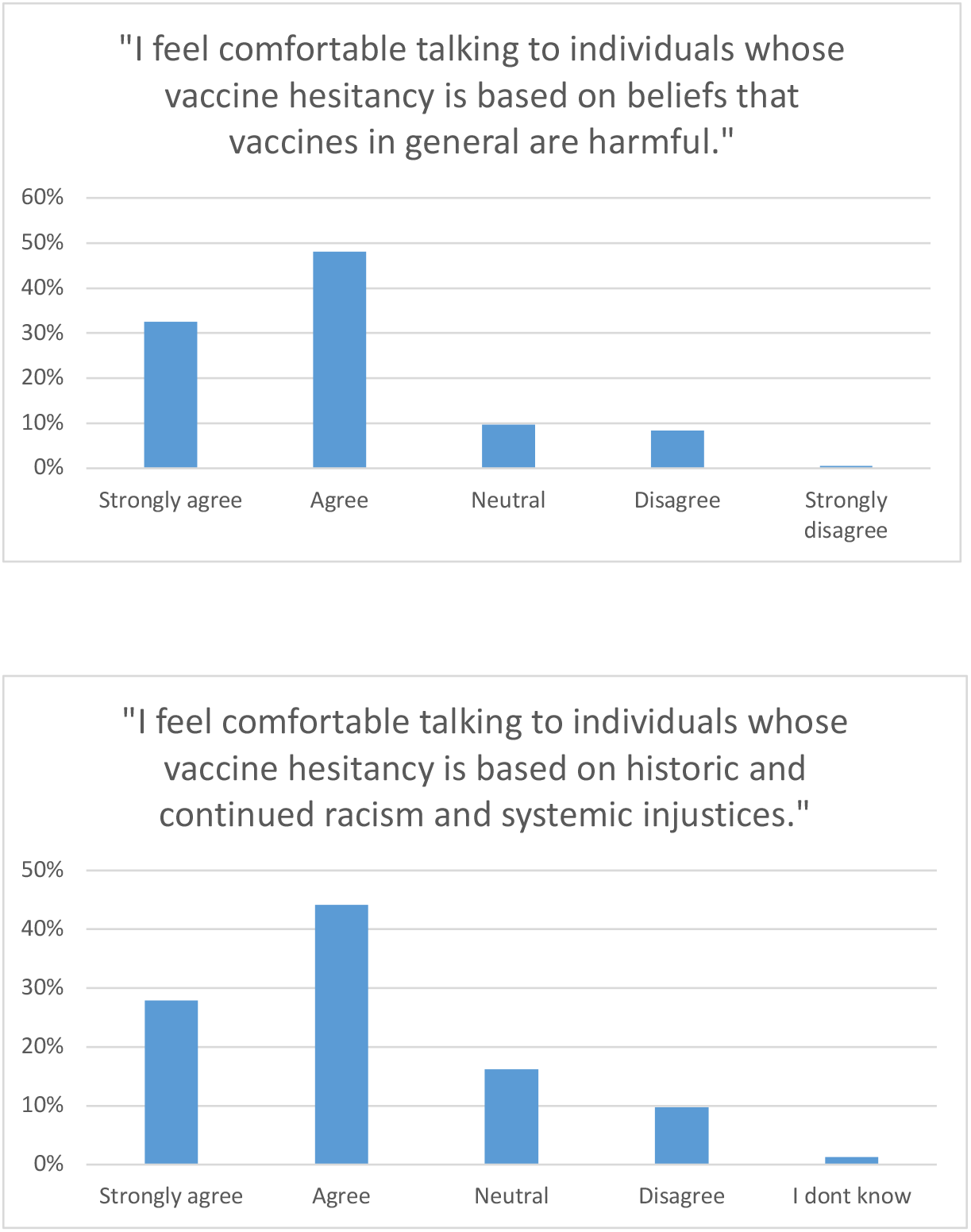
Provider Comfort with Counseling Patients with Vaccine Hesitancy

Most providers expressed some confidence in counseling individuals with vaccine hesitancy based on historic and continued racism and systemic injustices (111, 72.5%); however, only 43 providers (28.1%) reported feeling strongly confident with this counseling. Similarly, only 50 providers described the highest level of comfort counseling individuals with vaccine hesitancy grounded in general beliefs that vaccines are harmful. Those who believe the benefits of the vaccine outweigh the risks were also more likely to strongly agree with the statement that they feel comfortable talking to individuals whose vaccine hesitancy was based on historic and continued racism and systemic injustice (aOR 4.38, 95% CI 1.47-13.0) and to those whose hesitancy is based on beliefs that vaccines in general are harmful (aOR 5.56, 95% CI 1.98-15.6).

The sources that survey respondents most commonly used to find information regarding COVID-19 vaccination in pregnancy were the CDC (112 respondents, 74.2%), Hospital-specific resources (94 respondents, 62.3%) and ACOG (82 respondents, 54.3%). When asked to select the tool that would be most useful in helping them feel more confident in counseling pregnant patients regarding COVID-19 vaccination, 32 participants (20.9%) stated “A presentation from an expert in the field of obstetrics and gynecology”, 22 participants (14.4%) selected “A decision tool to use with my patients”, and 16 participants (10.5%) selected “A handout geared towards patients.”

## Discussion

Vaccination against COVID-19 is considered the cornerstone of pandemic management. CDC recommendations are permissive about COVID-19 vaccine use in pregnancy, though there is no explicit recommendation that pregnant people *should* receive the vaccine as there is for the general adult population. Guidance from the CDC and ACOG cite provider consultation as important (though not required) for decision-making about COVID-19 vaccination in pregnancy. Due to the absence of pregnancy specific vaccine evidence, and a lack of a clear directive to vaccinate all pregnant individuals, provider counseling will play a critical role in COVID-19 vaccine acceptance among pregnant people.

Provider understanding of and feelings towards vaccination have previously been shown to be key factors in their likelihood of recommending vaccination.^5-7^ Provider recommendation is also a significant factor for patient vaccine uptake.^8-11^ Despite the CDC’s offer for patients to discuss their decision with a healthcare provider and the pivotal role of healthcare providers in influencing vaccine uptake, little is known about how ready and confident providers feel to engage in this shared decision-making counseling.

In our study, respondents reported a high level of COVID-19 vaccine acceptance for themselves. Vaccinated healthcare providers have previously been shown to be more likely to recommend vaccines to their patients.^12^ The high levels of vaccine acceptance in this group suggests that it is a population that would be eager and amenable to counseling patients towards vaccination. Furthermore, most providers believe that the benefits of COVID-19 vaccination in pregnancy outweigh the risks, citing that pregnant individuals face increased risk of severe disease from COVID-19 infection and that there is low theoretical risk of harm from mRNA vaccine in pregnancy.

However, we identified a discrepancy between provider acceptance of vaccination as beneficial for pregnant individuals and their own comfort in engaging in conversations regarding the vaccine in pregnancy. Although 84.1% believed the benefits outweighed the risks in some or all circumstances, fewer expressed some confidence in answering questions about COVID-19 vaccine safety in pregnancy. Importantly, less than one-third of providers in our survey felt very confident in counseling pregnant patients about available evidence for mRNA vaccine safety in pregnancy, despite that nearly half had over 20 years of clinical experience. This disconnect between belief in efficacy and confidence in counseling is of particular concern and may impact the vaccination rates of patients

Overall, providers reported that they most trust information from their institution, the CDC, and ACOG regarding vaccination in pregnancy. Both ACOG and the CDC have developed tools for providers to guide conversations with patients around vaccination.^13-15^ These institutions can continue to provide guidance surrounding vaccine safety and efficacy as well as develop new counseling strategies for providers to improve comfort with counseling. They can engage community partners to involve those with vaccine hesitancy in vaccination campaigns and provider interactions. Based on our survey responses, these organizations may consider increased dissemination of information about ongoing presentations from experts in the field of obstetrics and gynecology and decision tools that providers may use with patients to improve confidence with vaccine counseling.

Although participants in our study reported moderate rates of confidence in counseling patients with vaccine hesitancy, less than one-third of participants strongly agreed that they felt confident in counseling patients with vaccine hesitancy based on historic and ongoing racism and systemic injustice. Provider lack of comfort in counseling those with vaccine hesitancy may further perpetuate racial inequities that have been highlighted by the pandemic, with disparities in vaccination rates as well as COVID-19 infection, hospitalization, and death rates.^16,17^ As others have pointed out, the concept of “vaccine hesitancy” puts the onus of disparities in vaccination rates on the patient rather than the provider and the healthcare systems that perpetuate mistrust and disparity.^18^ Our findings suggest a need for broader strategies for educating providers about the impact of systemic racism on both patient uncertainty as well as provider confidence in these encounters to improve confidence in vaccine counseling and ensuring equity in access. One long-term strategy proposed by Ojikutu et al is to longitudinally partner with and invest in communities of color from the initiation of vaccine research to community education, community-based organizations, and leadership in order to build trust and provide equitable inclusion in vaccination trials and implementation.^19^

Strengths of this study include that it represents a cross-sectional and inter-professional sample of specialties, practice settings, and provider types to surface the lived experiences of providers engaging in vaccine counseling for a vulnerable population during a pandemic. Limitations include that this study was performed in the early phases of vaccination; as more data surrounding vaccination in pregnancy become available, providers may have shifting comfort with this counseling. Furthermore, participants who opted to complete the survey may have a different level of confidence with this counseling compared to the general provider population. In addition, this sample represents a group of predominantly non-Hispanic White clinicians with high baseline vaccine trust, as nearly all providers received the vaccine themselves, and most stated that they always recommend the influenza and Tdap vaccines to pregnant patients. Nearly half spoke a second language, which may increase their confidence in counseling non-English speaking patients. As such, the confidence of this group may overrepresent the confidence of all clinicians who engage in vaccine counseling with pregnant patients. Finally, this study was conducted prior to the authorization of the vaccine developed by Janssen, which does not use an mRNA platform.

## CONCLUSION

As pregnant patients become increasingly eligible for COVID-19 vaccination nationwide, provider-initiated counseling regarding the safety and efficacy of vaccination will necessarily become a common occurrence in general medicine, primary care, and obstetric care. While the lack of inclusion of pregnant participants in the initial clinical trials of vaccination makes these discussions challenging, it also may increase their influence on vaccine uptake. Ensuring that providers feel comfortable bridging the gap between their belief that the vaccine is beneficial for pregnant patients and their comfort with holding conversations with patients regarding vaccination is paramount in order to ensure equitable access to vaccines for pregnant patients.

## Data Availability

All data produced in the present study are available upon reasonable request to the authors

## Notes

### Competing Interest Statement

The authors have declared no competing interest.

### Funding Statement

This study did not receive any funding

### Author Declarations

The study and survey instrument were approved by the Partners Healthcare IRB (IRB # 2021P000088).

